# Strong protective effect of the *APOL1* p.N264K variant against G2-associated focal segmental glomerulosclerosis and kidney disease

**DOI:** 10.1101/2023.08.02.23293554

**Authors:** Yask Gupta, David J. Friedman, Michelle McNulty, Atlas Khan, Brandon Lane, Chen Wang, Juntao Ke, Gina Jin, Benjamin Wooden, Andrea L. Knob, Tze Y. Lim, Gerald B. Appel, Kinsie Huggins, Lili Liu, Adele Mitrotti, Megan C. Stangl, Andrew Bomback, Rik Westland, Monica Bodria, Maddalena Marasa, Ning Shang, David J. Cohen, Russell J. Crew, William Morello, Pietro Canetta, Jai Radhakrishnan, Jeremiah Martino, Qingxue Liu, Wendy K. Chung, Angelica Espinoza, Yuan Luo, Wei-Qi Wei, Qiping Feng, Chunhua Weng, Yilu Fang, Iftikhar J. Kullo, Mohammadreza Naderian, Nita Limdi, Marguerite R. Irvin, Hemant Tiwari, Sumit Mohan, Maya Rao, Geoffrey Dube, Ninad S. Chaudhary, Orlando M. Gutiérrez, Suzanne E. Judd, Mary Cushman, Leslie A. Lange, Ethan M. Lange, Daniel L. Bivona, Miguel Verbitsky, Cheryl A. Winkler, Jeffrey B. Kopp, Dominick Santoriello, Ibrahim Batal, Sérgio Veloso Brant Pinheiro, Eduardo Araújo Oliveira, Ana Cristina Simoes e Silva, Isabella Pisani, Enrico Fiaccadori, Fangming Lin, Loreto Gesualdo, Antonio Amoroso, Gian Marco Ghiggeri, Vivette D. D’Agati, Riccardo Magistroni, Eimear E. Kenny, Ruth J.F. Loos, Giovanni Montini, Friedhelm Hildebrandt, Dirk S. Paul, Slavé Petrovski, David B. Goldstein, Matthias Kretzler, Rasheed Gbadegesin, Ali G. Gharavi, Krzysztof Kiryluk, Matthew G. Sampson, Martin R. Pollak, Simone Sanna-Cherchi

**Author notes:** **ADDRESS CORRESPONDENCE TO:** Simone Sanna-Cherchi, M.D., Division of Nephrology, Columbia University Vagelos College of Physicians and Surgeons, 1150 Street Nicholas Avenue, Russ Berrie Pavilion #412D, New York, New York 10032, USA.

## Abstract

Black Americans have a significantly higher risk of developing chronic kidney disease (CKD), especially focal segmental glomerulosclerosis (FSGS), than European Americans. Two coding variants (G1 and G2) in the *APOL1* gene play a major role in this disparity. While 13% of Black Americans carry the high-risk recessive genotypes, only a fraction of these individuals develops FSGS or kidney failure, indicating the involvement of additional disease modifiers.

Here, we show that the presence of the *APOL1* p.N264K missense variant, when co-inherited with the G2 *APOL1* risk allele, substantially reduces the penetrance of the G1G2 and G2G2 high-risk genotypes by rendering these genotypes low-risk. These results align with prior functional evidence showing that the p.N264K variant reduces the toxicity of the *APOL1* high-risk alleles. These findings have important implications for our understanding of the mechanisms of *APOL1*-associated nephropathy, as well as for the clinical management of individuals with high-risk genotypes that include the G2 allele.

## MAIN TEXT

Black Americans develop kidney disease at a rate five times higher than European Americans^1^. Two African ancestry-associated variants (G1 and G2) in the apolipoprotein L1 (*APOL1*) gene constitute major contributors to this disparity. *APOL1* is a component of the innate immune system targeting African trypanosomes, and the G1 and G2 variants likely rose to high population frequency by conferring resistance to *Trypanosoma brucei rhodesiense* (particularly G2) and *Trypanosoma brucei gambiense* (exclusively G1)^2,3^. However, the putative evolutionary benefits come at a cost of increased lifetime risk for kidney disease in individuals with two copies of these variants (i.e., G1/G1, G2/G2, or G1/G2). This is thought to be mediated by the ability of G1 and G2 variants to form cation-selective channels in podocytes resulting in subsequent activation of cytotoxic pathways^4-6^. This predisposes to progressive kidney disease, with odds ratios for hypertension-associated end stage kidney disease (ESKD), focal segmental glomerulosclerosis (FSGS), and HIV-associated nephropathy exceeding 7, 17, and 30, respectively, when comparing *APOL1* high-risk (*APOL1-HR*) to low-risk (*APOL1-LR*) genotypes^3,7^.

The number of at-risk individuals for *APOL1*-associated FSGS and kidney disease is considerable. In the United States, it is estimated that 13% of Black Americans carry two high-risk alleles^8^, and in certain West African populations, the rate of high-risk genotypes may be as high as 20-25%^8,9^. Approximately 15% of individuals with an *APOL1* high-risk genotype will develop ESKD, and a smaller fraction, estimated at 5%-8%, will develop FSGS^8^. Due to the high frequency of these genotypes, we estimate that at least 200,000 individuals in the US have *APOL1*-associated FSGS. The incomplete penetrance of *APOL1* high-risk genotypes is thought to reflect the requirement for disease modifiers that potentiate APOL1 cytotoxicity. A number of “second hits” have been proposed, with the most commonly recognized being high-interferon states (which result in increased APOL1 expression), either due to direct interferon administration, or caused by viral infections (e.g., HIV, SARS-CoV-2)^10,11^. Genetic modifiers have been suggested but, to date, the identification of modifier genetic variants for *APOL1*-mediated kidney disease and, particularly, FSGS, remains elusive. The few reported in the literature still require validation^12,13^.

In 2019, we studied the cytotoxic effect of multiple naturally and non-naturally occurring *APOL1* haplotypes in experimental cell-based systems. We found that the toxicity of G1 and G2 alleles was substantially reduced when expressed on the haplotype defined by the *APOL1* missense variant p.N264K (chr22:36265628 C>A; rs73885316)^14^, also associated with a partial loss of trypanolytic function^15^. These data suggested, at a functional level, a protective effect for this variant against the deleterious cellular effects of the G1 and G2 *APOL1* risk variants. The p.N264K defines one of the common G0 (non-risk) *APOL1* haplotypes, which is more frequent in individuals of European ancestry, but it is also present on a small fraction of G2 haplotypes in absence of G0, indicating two independent mutational events during evolution only on these two haplotypes. The p.N264K is therefore expected to be mutually exclusive with the APOL1 G1 allele.

To test the hypothesis that the G2-p.N264K haplotype differs in its genetic impact from the more common G2 risk allele without the p.N264K variant, we sought to compare its frequency in *APOL1-HR* subjects with FSGS to *APOL1-HR* controls without kidney disease (**Figure 1A**). First, to eliminate potential confounding by the p.N264K haplotype defined by the more common *APOL1* non-risk G0 allele, we excluded all individuals with non-risk, G0-containing genotypes, i.e., G0/G0, G0/G1, and G0/G2. We studied two case-control FSGS discovery cohorts: the first consisted of 434 *APOL1-HR* FSGS cases and 2,398 genetically matched *APOL1-HR* population controls subjected to Illumina DNA microarray genotyping and imputation; the second included 94 *APOL1-HR* FSGS cases and 208 genetically matched *APOL1-HR* controls with whole genome sequencing data (**Supplementary Figure S1**), for a total of 528 FSGS cases and 2,606 population controls with no known kidney disease. Next, in order to investigate the impact of the p.N264K variant, we conducted a comprehensive analysis only on *APOL1* high-risk individuals, employing categorical approaches (based on allelic frequency) and, as sensitivity analysis, regression-based (based on genotypes) statistical tests. The primary analysis was conducted on categorical variables using a Cochran– Mantel–Haenszel (CMH) test and considering potential confounding factors such as sex and array-based vs sequence-based genotyping. We then conducted a set of sensitivity analyses: first, we used Firth’s regression test and also incorporated principal components (PCs) as covariates in order to account for potential residual population stratification; second we conducted haplotype-of-origin analysis using Tractor^16^.

**Figure 1.**
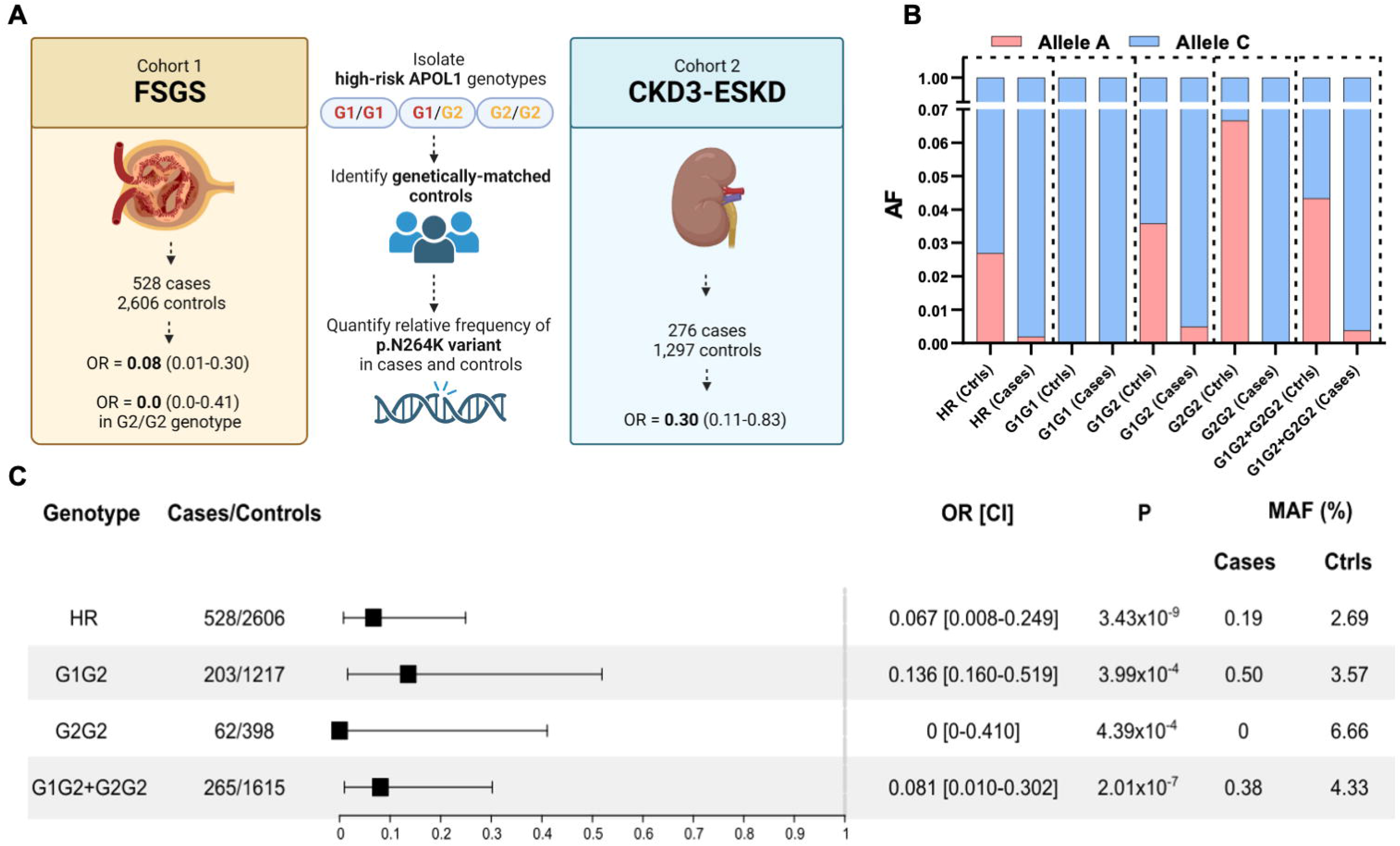
Protective effect of the *APOL1* p.N264K missense variant against G2-associated FSGS. **A)** Graphical representation of the study design, cohorts and main results of the study. Stratified association analysis of the combined cohort of 528 *APOL1* high-risk FSGS and 2,606 genetically-matched *APOL1* high-risk controls: **B)** stacked bar plot for the p.N264K MAF across *APOL1*-HR genotypes in cases and controls; the Allele C is the reference allele encoding for the p.N264; the Allele A is the minor allele resulting the p.K264 variant aminoacid; **C)** Forest plot for the p.N264K association analysis showing significantly protective odds ratios across *APOL1-HR* genotypes. FSGS=focal segmental glomerulosclerosis; CKD=chronic kidney disease; ESKD=end-stage kidney disease; AF=allele frequency; Ctrls=controls; OR=odds ratio; CI=95% confidence interval; MAF=minor allele frequency.

In our *APOL1-HR* FSGS cohorts, we observed a strong protective effect for the p.N264K minor allele ‘A’ (MAF cases = 0.19 % and MAF controls = 2.7%, OR=0.07, 95%CI = 0.01-0.25, CMH test *P*=3.4×10^−9^) as compared to *APOL1-HR* controls. Stratifying the cohort for the three *APOL1* high-risk genotypes showed that this variant was only observed within *APOL1-HR* individuals carrying the G2 allele (i.e., G1/G2 and G2/G2) and, as expected, never in G1/G1 subjects (**Figure 1, Supplementary Figure S2**). These findings support a protective effect of the p.N264K variant only in the context of G2-containing *APOL1-HR* genotypes. In fact, the p.N264K variant seemed to confer complete protection against FSGS as it was never observed in cases in the presence of the G2/G2 genotype: OR=0, 95%CI 0-0.41; CMH test *P*=4.4×10^−4^. A strong and significant protective effect was also observed for the G1/G2 genotype with a p.N264K MAF of 3.57% in controls as compared to 0.49% in cases (OR=0.14, 95%CI:0.16-0.52; CMH test *P*=4.0×10^−4^). Consistent with these findings, analyzing individuals with G1/G2 or G2/G2 genotypes combined increased the level of statistical significance for the p.N264K protective effect (OR=0.08, 95%CI 0.01-0.3, CMH test *P*=2×10^−7^).

The FSGS case-control samples were well-matched on principal component analysis (PCA) (**Supplementary Figure S1**). In fact, in our sensitivity analyses that additionally adjust for population structure confirmed the results obtained by CMH, as Firth’s regression tests confirmed the strong protective effect of the p.N264K variant against FSGS with comparable effect sizes (**Supplementary Figure S2**).

As expected from population distribution of haplotypes, in the context of *APOL1-HR* genotypes, the p.N264K is limited to G2-containing genotypes (i.e., G1/G2 or G2/G2). Nevertheless, a recombination event between the p.N264K and the G1 or G2 alleles (although very unlikely given the proximity of these APOL1 alleles), could result in contamination from the European G0-p.N264K haplotype due to local ancestry admixture. To finally exclude this scenario, in our final sensitivity analysis we conducted haplotype-of-origin analysis in the discovery cohort using Tractor^16^, a statistical framework that deconvolutes the local haplotypes into ancestral (in this case European and African) haplotypes. This confirmatory analysis showed a significant protective effect of the p.N264K variant exclusively originating from the African haplotype (OR=0.10, 95%CI=0.02-0.29, *P*=1.3×10^−7^), while the European haplotype was non-significant (OR_(ADJ)_=0.74, 95%CI= 0.00-11.37, *P*=0.85) despite larger sample size (**Supplementary Figure S3**). Again, stratifying for G1/G2 or G2/G2 further validated the G2-specific protective effect of the p.N264K variant for the African haplotype (OR=0.12, 95%CI=0.02-0.35, *P*=3.53×10^−6^) but not for the European haplotype (OR_(ADJ)_=0.76, CI=0.00-12.75, *P*=0.86).

Overall, these results support a strong protective effect of the *APOL1* p.N264K missense variant against *APOL1*-associated FSGS, but this effect occurs exclusively on G2-containing *APOL1* high-risk genotypes of African origin. In practical terms, based on these analyses, *APOL1-HR* individuals are at least 8.3 times less likely to develop FSGS if they carry one copy of the p.N264K missense variant.

Finally, to test the generalizability of these findings to milder forms of *APOL1*-associated kidney disease, we investigated the protective effect of the *APOL1* p.N264K in individuals from the REasons for Geographic and Racial Differences in Stroke (REGARDS)^17^ and Electronic Medical Records and Genomics Phase III (eMERGE-III)^18^ studies. In aggregate, these cohorts included 1,573 *APOL1-HR* individuals with available kidney function data. Of these, 276 had CKD stage 3 or worse (considered as cases), and 1,297 genetically-matched *APOL1-HR* controls with estimated glomerular filtration rate (eGFR) >60ml/min/1.73m^2^ (**Supplementary Figure S4**). Despite the smaller sample size, milder form of *APOL1*-associated kidney disease, and incomplete clinical data to classify and exclude unrelated causes for CKD in these cohorts, the findings revealed a direction-consistent protective effect for the p.N264K variant among individuals with the *G2-APOL1-HR* genotypes, by which p,N264K carriers were 3.3 times less likely to have CKD3 or worse (OR=0.30, 95%CI: 0.11-0.83, CMH *P*=0.023, **Supplementary Table S2**), with this likely representing an underestimation due to confounders as mentioned above.

In conclusion, here we report on the strong protective effect of the *APOL1* p.N264K missense variant against G2-mediated FSGS and kidney disease. These results have immediate and broad implications for translational research and clinical practice. First, from the genetic standpoint, it is important to note that we observed a very large effect of p.N264K on mitigating the consequences of the G2 risk allele but saw no evidence of this variant on the more common G1 risk allele. As consequence, because p.N264K and G1 alleles are mutually exclusive, this finding raises the possibility of additional genetic modifiers specific to G1 and, in general, identifiable by considering genotype-specific *APOL1* studies. In addition to studies of the *APOL1* high-risk genotype as a single genetic driver, analyses conducted by partitioning cohorts into the three specific *APOL1* high-risk genotypes, although might require larger sample sizes, are likely to provide significant additional insight into the genetics and underlying biology of *APOL1*-associated FSGS and kidney disease. Second, our genetic observations are in agreement with our previous functional studies showing that the p.N264K variant is able to reverse the cytotoxic effect of both G1 and G2 risk variants in cell-based assays^14^.

Taken together, these data support the hypothesis that the p.N264K missense variant negates the toxic effect of the G2 allele, and will allow the reclassification of a fraction of *APOL1* G1/G2 or G2/G2 high-risk individuals as having a non-high-risk genotype if p.N264K is also present. This discovery has substantial, immediate, and clinically-relevant implications. First, individuals affected by CKD or ESKD with *APOL1* G1/G2 or G2/2 high-risk genotypes but with the p.N264K missense variant are unlikely to have APOL1-associated FSGS, and therefore an additional cause (immune, toxic, structural, or others) should be investigated because this will likely result in a different therapeutic approach. Second, importantly, in kidney transplant settings, these results can significantly affect donor selection and both donor kidney, and recipient graft, outcome. In fact, *APOL1* G2-HR donors who are p.N264K positive will likely have kidney outcomes similar to any of the G1G0, G2G0, and G0G0 low-risk donors, thus expanding donors’ pool; kidney transplant recipients of a *APOL1*-HR-p.N264K kidney will likely have low risk for developing *de novo* FSGS on the graft or graft failure from APOL1-associated kidney disease. Third, incorporation of this knowledge will allow more accurate study design for new intervention trials by which individuals with *APOL1*-HR-p.N264K genotypes should not be included in the intervention arm as cases since this genotype is genetically and functionally a low-risk genotype. Finally, the knowledge presented here will affect family risk stratification and planning, and, in general, CKD risk ascertainment at the population level.

## Data Availability

All data produced in the present study are available upon reasonable request to the authors

## ACKNOWLEDGEMENTS

We thank the patients and their family members for participating in this study.

