## Supplementary Appendix for "Strong protective effect of the *APOL1* p.N264K variant against G2-associated focal segmental glomerulosclerosis and kidney disease"

**SUPPLEMENTARY METHODS**

**FSGS cohorts, controls, genotyping and imputation, and association tests**

*FSGS case-control cohort 1*: The cohort consisted of 434 FSGS *APOL1-HR* cases (**Supplementary Table S1**) and 2,398 *APOL1-HR* controls. The genotyping of the cases was performed using multiple versions of the Illumina MEGA (Multi-Ethnic Global Array) chips (n = 196) that included MEGA 1.0, MEGA 1.1 and MEGA^EX^, and the Illumina HumanOmniExpress-12 (n = 238). The controls were genotyped on MEGA1.0 and were selected based on genetic ancestry and *APOL1* genotype status from over 50,000 individuals from the PAGE consortium^1^. We extracted G1 (rs73885319) and G2 (rs71785313) from the MEGA arrays to define *APOL1* high-risk cohort. Also, the p.N264K (chr22:36265628 C>A; rs73885316) variant was included on the MEGA arrays and hence directly genotyped, while imputed with R2 >0.8 in the Illumina HumanOmniExpress-12. The differences between the chips were corrected first by mapping all the SNPs to a common cluster file in Genome Studio software for individual platforms and further using Snpflip (https://github.com/biocore-ntnu/snpflip) software. In total, we used 767,100 SNPs as input for imputation after quality control, which included filtration for MAF > 1%, missing SNPs < 95%, HWE (controls) *P* < 0.00001, and Macarthy Group tools for strand bias and removal of SNPs that deviated from expected allele frequency using the 1000 Genome Project. The same quality control was applied for cases from HumanOmniExpress-12 separately.

We performed imputation on *APOL1-HR* cases (MEGA and HumanOmniExpress) and controls (MEGA) together using the TopMed reference imputation panel. SNPs with R2 > 0.8, MAF > 1%, missing SNPs < 95%, and HWE (controls) P > 0.00001 were retained. All analyses were done on unrelated samples after removing the relatedness up to two degree using KING2.3.0. PCs were calculated using PLINK 2 based on the LD-pruned SNPs.

*FSGS case-control cohort 2*: The cohort for this study consisted of 94 *APOL1-HR* cases (refer to Supplementary Table S1) and 208 *APOL1-HR* controls, all subjected to 30X whole-genome sequencing (WGS). To obtain the genetic data, the raw FASTQ files for the cases were aligned to the hg19 assembly. The alignment data underwent processing using the DRAGEN pipeline, resulting in recalibrated GVCFs (Genomic VCFs). The GVCFs were jointly called using GATK 4.3 separately for samples from the DUKE and CureGN cohorts. For the control group, we included *APOL1-HR* samples obtained from the 1000 Genomes Project, MESA cohort, and internal controls from the Columbia University Institute for Genomic Medicine (IGM). Genotypes were extracted after performing internal harmonization. Initially, all the case samples were lifted from the hg19 to the hg38 assembly using the rtracklayer R package. Then, the genotypes from the cases and controls were merged based on common SNPs.

The *APOL1* G1, G2, and p.N264K variants were directly sequenced to obtain specific genetic information. To ensure the analysis was performed on unrelated individuals, we removed relatedness up to two degrees using the KING2.3 software. The same quality control measures were applied to the FSGS cohort 2, as for the initial FSGS cohort, before calculating principal components (PCs).

**Statistical analysis**

Stratified analysis on alleles was conducted on the two FSGS cohorts using the Cochran-Mantel-Haenszel (CMH) test statistic. The CMH test was performed using the mantelhaen.test function in R with the exact=TRUE option. The cohorts were stratified based on the variables of cohort and sex for each of the haplotypes (G1/G1, G1/G2, G2/G2) both individually and combined.

Furthermore, Firth regression was performed separately for each haplotype within each cohort using PLINK2. The covariates used in the regression analysis were sex and the first two principal components (PCs). Finally, a meta-analysis was conducted to combine the results from the two cohorts. The fixed-effect model was utilized, considering the effect sizes and standard errors obtained from the PLINK2 analysis.

**Ancestry resolution analysis at the *APOL1* locus on FSGS cohort 1**

Phased genotypes from the first FSGS cohort were utilized after imputation with the TOPMed reference panel. These genotypes were employed for predicting local ancestry inference (LAI) using RFMix v2. The LAI prediction was performed against samples harboring common variants obtained from the 1000 Genomes Project, specifically YRI (representing the African population) and CEU (representing the European population). The output from RFMix was subsequently integrated into the Tractor pipeline to deconvolute ancestry-specific dosages, variant call files (VCFs), and haplotype counts for each sample and SNP^2^. Within Tractor, ancestry-specific haplotype counts and dosages for the p.N264K variant were extracted. Firth regression analysis was conducted using the logistf R package, incorporating sex, admixture fraction (derived from RFMix), and haplotype counts as covariates in a fixed effect model. This analytical approach facilitated the assessment of the p.N264K variant's association with FSGS by incorporating local ancestry inference, deconvolution of dosages and haplotype counts, and regression analysis with appropriate covariates.

**CKD cohorts, genotyping, and analysis**

*The REGARDS study*: The REGARDS study investigates the incidence of stroke in a population of 30,239 Black and White adults (≥45 years of age)^3^. Within this study, we identified 8,198 Black participants with genotyped *APOL1* risk alleles (G1 & G2) using TaqMan SNP Genotyping Assay ^4^ and genome-wide genotyping using the Multi-Ethnic Genotyping Array (MEGA). To increase sample size, we imputed *APOL1* genotypes for an additional 534 subjects using the TOPMed Imputation Server ^5^. Using kinship analysis, we identified and removed related samples (up to 2nd degree) between the REGARDS and PAGE consortiums. Finally, we removed all individuals with an APOL1 low-risk genotype, i.e. G0/G0, G0/G1, G0/G2. Our final cohort was composed of 1,043 *APOL1* high-risk individuals with genotypes G1/G1 (n=417), G1/G2 (n=455) and G2/G2 (n=171). The p.N264K variant was included and directly genotyped in the MEGA array. To compare the allele frequency of the p.N264K variant, we stratified samples into the two following groups: cases - estimated glomerular filtration rate (eGFR) < 60, or ESKD, or self-reported kidney failure; control: eGFR > 60. eGFR was measured from the CKD-Epi equation.

*Electronic Medical Records and Genomics (eMERGE) study*: The eMERGE network has made available electronic health record (EHR) information connected to GWAS data for a total of 102,138 individuals^6^. These individuals were recruited in three phases (eMERGE III) from 12 participating medical centers spanning the years 2007 to 2019. The study cohort consisted of 54% females, with an average age of 69 years. Self-reported demographics indicated that 76% identified as European, 15% as African American, 6% as Latinx, and 1% as East or Southeast Asian.

Every individual underwent genome-wide genotyping, and the specific procedures for genotyping, quality control analyses, and imputation have been previously documented^7^. Principal components (PCs) were derived using FlashPCA^8^ on a collection of 48,509 common variants (minor allele frequency ≥ 0.01) that were independent (pruned in PLINK using the --indep-pairwise 500 50 0.05 command). Imputation of the APOL1 variants G1 (R2 = 0.998), G2 (R2 = 0.995), and N264K (R2 = 0.8445) was performed using the TOPMed imputation server, as outlined in the referenced publication^9^. From this cohort, we selected 530 *APOL1*-HR individuals with available kidney function data in order to classify 126 cases based on eGFR <60 (CKD3-5 or ESKD) and 404 controls (eGFR >60), in the same way as for the REGARDS study above.

**IRB approval**

Written informed consent was collected from all participating patients seen at Columbia (and collaborating Institutions) and/or their guardians in accordance with the Columbia University Institutional Review Board (Protocol AAAC7385) and the policy on bioethics and human biologic samples of AstraZeneca. All internationally recruited patients and/or their guardians were consented according to the Declaration of Helsinki and in compliance with the local ethic committees, as part of the parent IRB protocol approved at Columbia University.

**SUPPLEMENTARY FIGURES**

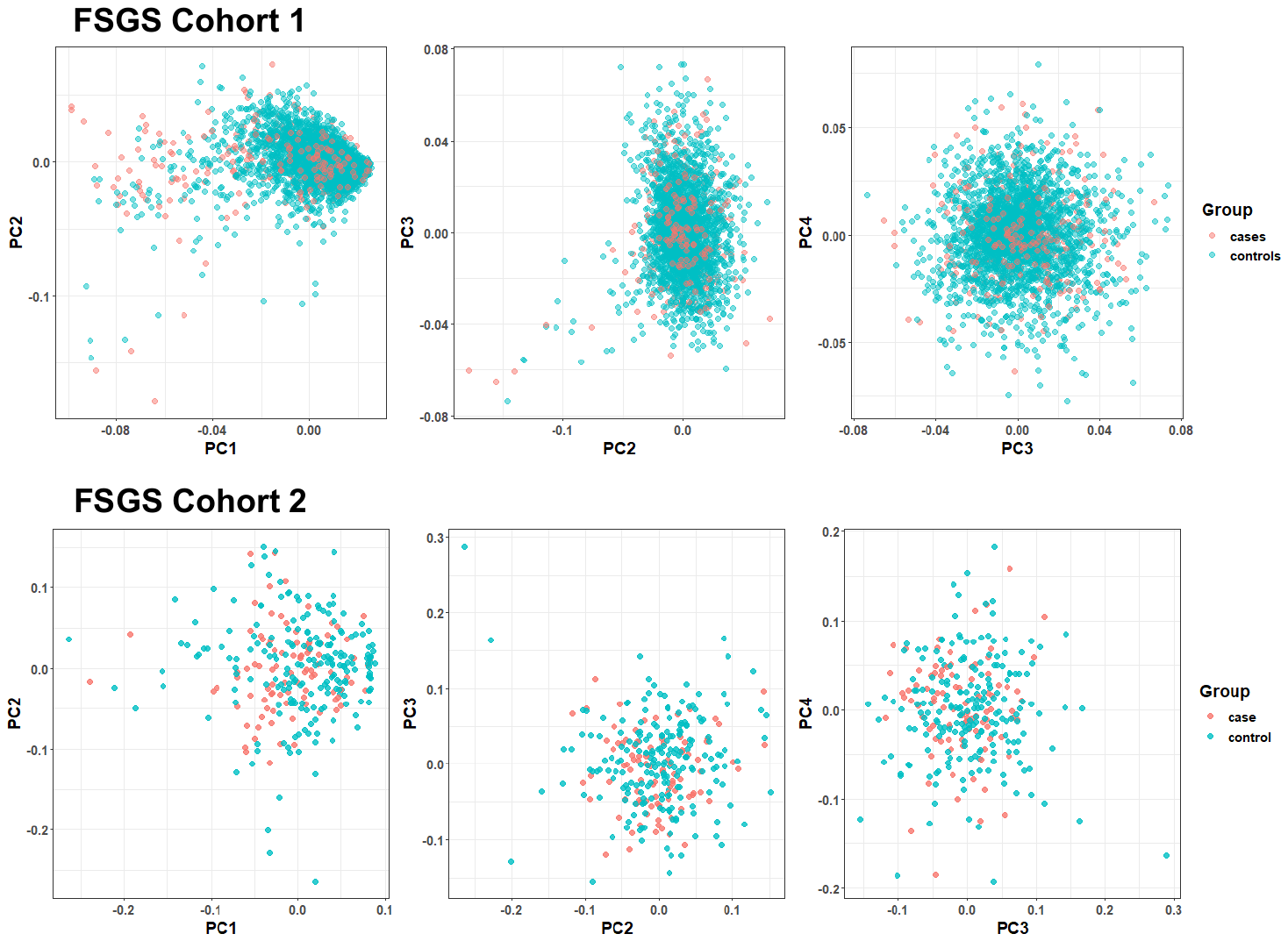
**Supplementary Figure S1. Principal component analysis of FSGS cohorts.**

The figure shows plots for the first three principal components for the FSGS cohort 1 based on 279,502 pruned SNPs and for the FSGS cohort 2 based on 219,146 pruned SNPs. PCA shows well-matched case-control data for both cohorts.

PC=principal component.

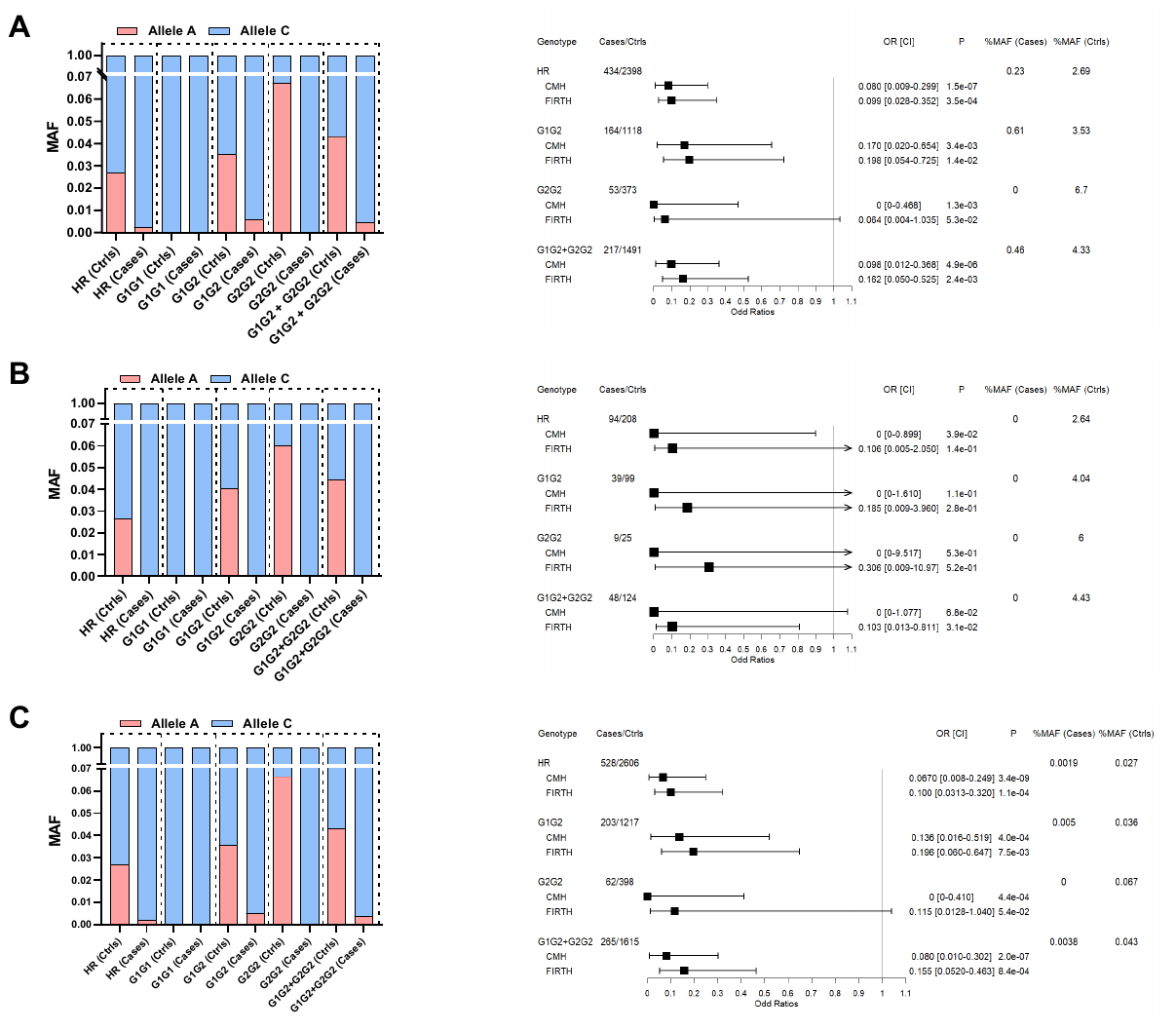
**Supplementary Figure S2. *APOL1* p.N264K association analysis in FSGS cohorts 1 and 2.**

The figure shows stacked bar plots on the left panels and forest plots on the right panels for **A)** FSGS cohort 1, **B)** FSGS cohort 2 **C)** Meta-analysis of the pooled cohort. The right panel shows forest plots for all cohorts with *APOL1* genotypes in the first column, number of cases and controls in the second column, odd ratios (pooled for CMH and adjusted odd ratios for firth regression) and confidence interval graph in the third column followed by *P* value, percentage of minor allele in cases and controls. For every *APOL1* HR genotype the forest plot also provides summary statistics for both the CMH test and firth regression. CI=95% confidence interval.

**Supplementary Figure S3. *APOL1* p.N264K association with FSGS in EUR and AFR population.**

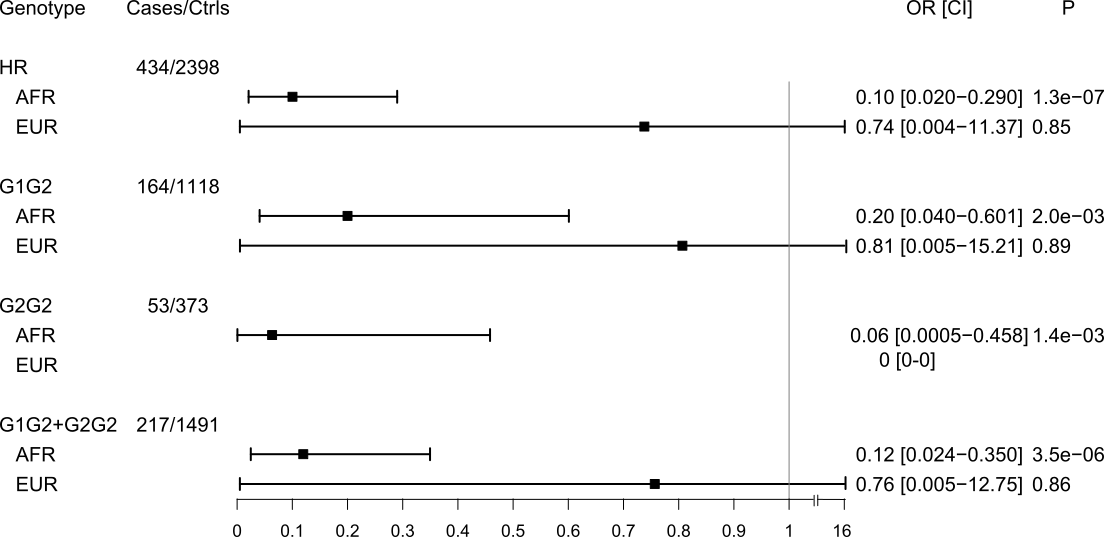

The forest plot describes the summary of local ancestry statistics using Tractor^2^. Every group of row shows different *APOL1-HR* genotypes and, in sub rows, the individual local ancestry (AFR, EUR) association after estimation with RFMix. The plot shows adjusted odd-ratios and confidence intervals obtained using firth regression after adjustment for sex and two PCs. The *P* values are provided in last column.

**Supplementary Figure S4. Principal component plot of *APOL1-HR* participants in the REGARDS and EMERGE-III Studies**

A.
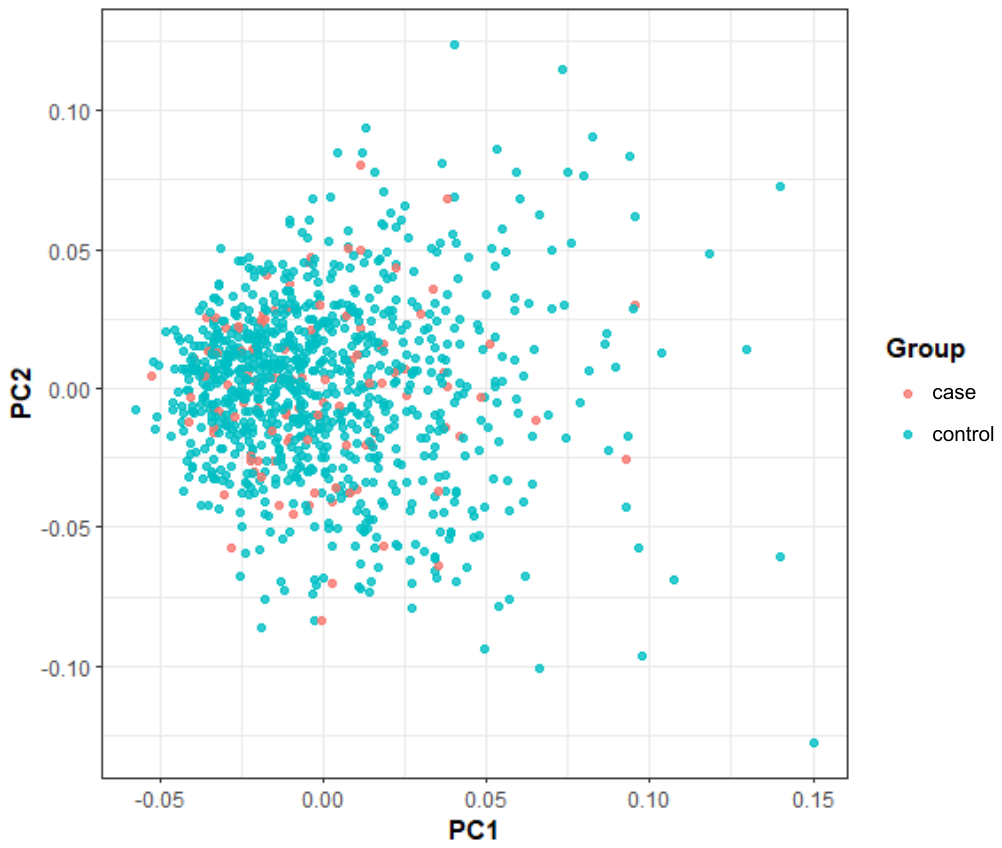
REGARDS
B.
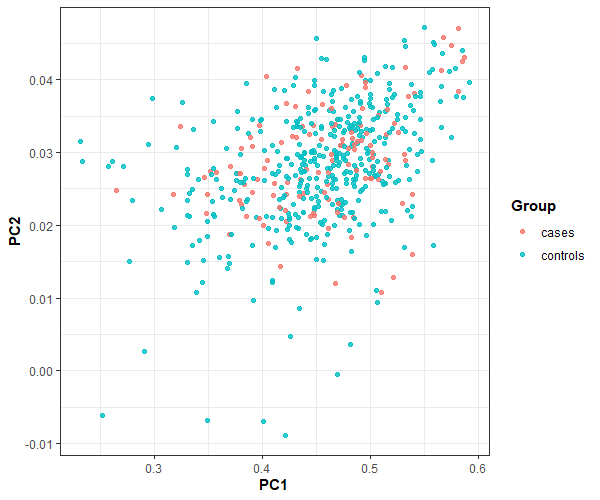
eMERGE-III

**SUPPLEMENTARY TABLES**

**Supplementary Table S1. FSGS case-control discovery and replication cohorts**

| **FSGS cohort 1** |  |  |
| --- | --- | --- |
| **Recruitment Center** | **Number of cases** | **Genotyping Platform** |
| CUIMC | 196 | Illumina MEGA arrays |
| BIDMC | 238 | Illumina HumanOmniExpress-12 |
| **Total** | **434** |  |
| **FSGS cohort 2** |  |  |
| **Recruitment Center** | **Number of cases** | **Genotyping Platform** |
| CureGN + Neptune + CUIMC | 58 | 30X WGS |
| Duke University | 36 | 30X WGS |
| **Total** | 94 |  |

**Supplementary Table S2. Association of *APOL1* p.N264K to CKD-5 and ESKD in APOL1-HR individuals from the REGARDs and EMERGE-III studies.**

| **REGARDS** |  |  |  |  |  |  |  |  |  |
| --- | --- | --- | --- | --- | --- | --- | --- | --- | --- |
| **APOL1 GT** | **MAF (%)** | **MAF CA (%)** | **MAF CO (%)** | **N. CA** | **N. CO** | **Fet OR** | **L5** | **U5** | **Fet P** |
| HR | 2.30 | 0.67 | 2.58 | 150 | 893 | 0.261 | 0.030 | 1.006 | 0.055 |
| G1G2 | 2.64 | 0.73 | 2.97 | 68 | 387 | 0.242 | 0.006 | 1.517 | 0.239 |
| G2G2 | 7.02 | 2.23 | 7.72 | 22 | 149 | 0.279 | 0.007 | 1.810 | 0.338 |
| G1G2+G2G2 | 3.83 | 1.11 | 4.29 | 80 | 536 | 0.251 | 0.029 | 0.0.974 | 0.036 |
| **EMERGE-III** |  |  |  |  |  |  |  |  |  |
| **APOL1 GT** | **MAF (%)** | **MAF CA (%)** | **MAF CO (%)** | **N. CA** | **N. CO** | **Fet OR** | **L5** | **U5** | **Fet P** |
| HR | 1.98 | 0.79 | 2.35 | 126 | 404 | 0.332 | 0.037 | 1.394 | 0.192 |
| G1G2 | 2.10 | 0.91 | 2.46 | 55 | 183 | 0.364 | 0.008 | 2.681 | 0.466 |
| G2G2 | 8.59 | 5.55 | 9.09 | 9 | 55 | 0.590 | 0.013 | 4.660 | 1 |
| G1G2+G2G2 | 3.48 | 1.56 | 3.99 | 64 | 238 | 0.382 | 0.043 | 1.619 | 0.276 |
| **JOINT REGARDS – EMERGE-III** | | | | | | | | | |
| **APOL1 GT** | **MAF (%)** | **MAF CA (%)** | **MAF CO (%)** | **N. CA** | **N. CO** | **CMH OR** | **L5** | **U5** | **CMH P** |
| HR | 2.19 | 0.72 | 2.51 | 276 | 1297 | 0.286 | 0.103 | 0.792 | 0.016 |
| G1G2 | 2.44 | 0.81 | 2.81 | 123 | 570 | 0.290 | 0.033 | 1.155 | 0.103 |
| G2G2 | 7.45 | 3.22 | 8.09 | 31 | 204 | 0.378 | 0.043 | 1.544 | 0.295 |
| G1G2+G2G2 | 3.71 | 1.29 | 4.19 | 144 | 774 | 0.300 | 0.108 | 0.834 | 0.023 |

GT=genotype; MAF=minor allele frequency for the p.N264K variant; CA=cases; CO=controls; N.=number; Fet=Fisher’s Exact Test; L5=lower 5% confidence interval; U5=upper 5% confidence interval.
